# Neural network-derived electrocardiographic features have prognostic significance and important phenotypic and genotypic associations

**DOI:** 10.1101/2023.06.15.23291428

**Authors:** Arunashis Sau, Antonio H. Ribeiro, Kathryn A. McGurk, Libor Pastika, Nikesh Bajaj, Maddalena Ardissino, Jun Yu Chen, Huiyi Wu, Xili Shi, Katerina Hnatkova, Sean Zheng, Annie Britton, Martin Shipley, Irena Andršová, Tomáš Novotný, Ester Sabino, Luana Giatti, Sandhi M Barreto, Jonathan W. Waks, Daniel B. Kramer, Danilo Mandic, Nicholas S. Peters, Declan P. O’Regan, Marek Malik, James S. Ware, Antonio Luiz P. Ribeiro, Fu Siong Ng

**Affiliations:** National Heart and Lung Institute, Imperial College London, United Kingdom; Department of Cardiology, Imperial College Healthcare NHS Trust, London, United Kingdom; Department of Information Technology, Uppsala University, Uppsala, Sweden; Research Department of Epidemiology and Public Health, University College London, United Kingdom; Department of Internal Medicine and Cardiology, University Hospital Brno and Masaryk University, Brno, Czech Republic; Department of Infectious Diseases, School of Medicine and Institute of Tropical Medicine, University of São Paulo, São Paulo, Brazil; Department of Preventive Medicine, School of Medicine, and Hospital das Clínicas/EBSERH, Universidade Federal de Minas Gerais; Harvard-Thorndike Electrophysiology Institute, Beth Israel Deaconess Medical Center, Harvard Medical School, Boston, MA, USA; Richard A. and Susan F. Smith Center for Outcomes Research in Cardiology, Beth Israel Deaconess Medical Center, Harvard Medical School, Boston MA USA; Department of Electrical and Electronic Engineering, Imperial College London, United Kingdom; MRC London Institute of Medical Sciences, Imperial College London, London, UK; Royal Brompton & Harefield Hospitals, Guy’s and St. Thomas’ NHS Foundation Trust, London, UK; Department of Internal Medicine, Faculdade de Medicina, and Telehealth Center and Cardiology Service, Hospital das Clínicas, Universidade Federal de Minas Gerais, Belo Horizonte, Brazil

**Keywords:** machine learning, neural network, phenogroup, risk prediction, electrocardiogram

## Abstract

**Background:** Subtle prognostically-important ECG features may not be apparent to physicians. In the course of supervised machine learning (ML), many thousands of ECG features are identified. These are not limited to conventional ECG parameters and morphology.

**Hypothesis:** Novel neural network (NN)-derived ECG features can predict future cardiovascular disease and mortality

**Methods and Results:** We extracted 5120 NN-derived ECG features from an AI-ECG model trained for six simple diagnoses and applied unsupervised machine learning to identify three phenogroups. In the derivation cohort (CODE, 1,558,421 subjects), the three phenogroups had significantly different mortality profiles. After adjusting for known covariates, phenogroup B had a 20% increase in long-term mortality compared to phenogroup A (HR 1.20, 95% CI 1.17-1.23, p < 0.0001). The predictive ability of the phenogroups was retained in a group with physician confirmed normal ECGs. We externally validated our findings in five diverse cohorts (Figure) and found phenogroup B had a significantly greater risk of mortality in all cohorts. Phenome-wide association study (PheWAS) showed phenogroup B had a higher rate of future AF, ischaemic heart disease, AV block, heart failure, VT, and cardiac arrest.

Phenogroup B had increased cardiac chamber volumes and decreased cardiac output. A single-trait GWAS yielded four loci. SCN10A, SCN5A and CAV1 have roles in cardiac conduction and arrhythmia. ARHGAP24 does not have a clear cardiac role and may be a novel target. Gradient-weighted Class Activation Mapping (Grad-CAM) identified the terminal QRS and terminal T wave as important regions of the ECG for identification of phenogroup B.

**Conclusion:** NN-derived ECG features can be used to predict all-cause mortality and future cardiovascular diseases. We have identified biologically plausible and novel phenotypic and genotypic associations that describe mechanisms for the increased risk identified.

## Introduction

The electrocardiogram (ECG) is a widely used investigation for the assessment of cardiovascular disease. The ECG captures information relating to a wide range of cardiac pathology, from cardiomyopathy and channelopathies to conduction system disease. Several specific ECG features have been associated with adverse long-term prognosis, including left bundle branch block (LBBB) (1), prolongation of QRS duration (2), and ECG changes consistent with left ventricular hypertrophy (3). However, these crude ECG features are based on physician interpretation or ECG measurements and there may be other ECG features, not easily discernible by humans, that may be as clinically and prognostically meaningful.

Recently, there has been a rapid increase in applications of artificial intelligence (AI) to the analysis of the ECG (4, 5, 6). Supervised machine learning (ML) is the most common form of machine learning applied to ECGs, where a model is trained to predict a label e.g., future atrial fibrillation or impaired left ventricular function (5, 6). AI-ECG models often use a convolutional neural network (CNN) architecture and have generally performed well (4, 5, 6). Although the classification task is often specific and singular, the CNN identifies many thousands of ECG features over the course of model training, which are not limited to conventional ECG parameters and morphology, and applies these features to make the label prediction (7). These novel neural network (NN)-derived ECG features have not been extensively studied. There is the potential that they may be universal, transferable, and applicable to a range of clinically useful tasks.

We hypothesised that the ECG features learned by a neural network may have implications beyond the original task for which it was trained and may have important clinical, phenotypic, and genotypic associations. We applied an existing AI-ECG model trained to classify six common ECG diagnoses relating to rhythm and conduction disease (8). We extracted the NN-derived ECG features from the penultimate layer of the CNN and applied unsupervised machine learning to identify clinically distinct phenogroups with prognostic significance. Given the critical role of external validation (9), we validated our analysis in four external datasets across two continents with diverse ancestral backgrounds. Furthermore, our model was evaluated across diverse cohorts including volunteers, primary care patients and patients with established cardiomyopathy. We subsequently explored potential biological mechanisms underlying the associations of the ECG phenogroups with survival by evaluating their associations with a wide range of phenotypes and genetics. Finally, we performed Mendelian randomisation (MR) to explore bidirectional associations of the genetically predicted ECG phenogroups with cardiovascular traits and outcomes.

## Methods

### Ethical approvals

This study complies with all relevant ethical regulations. The Clinical Outcomes in Digital Electrocardiography (CODE) study was approved by the Research Ethics Committee of the Universidade Federal de Minas Gerais, protocol 49368496317.7.0000.5149. The Whitehall II study was approved by the Joint University College London/University College London Hospitals Committees on the Ethics of Human Research. The UK Biobank has approval from the North West Multi-Centre Research Ethics Committee as a Research Tissue Bank (application ID 48666). The Longitudinal Study of Adult Health (ELSA-Brasil) was approved by the Research Ethics Committees of the participating institutions and by the National Committee for Research Ethics (CONEP 976/2006) of the Ministry of Health. The São Paulo-Minas Gerais Tropical Medicine Research Center (SaMi-Trop) study was approved by the Brazilian National Institutional Review Board (CONEP), No. 179.685/2012. For the Beth Israel Deaconess Medical Center (BIDMC) cohort ethics review and approval was provided by the Beth Israel Deaconess Medical Center Committee on Clinical Investigations, IRB protocol # 2023P000042.

### ECG datasets

#### (i) The CODE Cohort

The CODE cohort is a database of 2,322,513 ECG records from 1,676,384 different patients of 811 counties in the state of Minas Gerais/Brazil from the Telehealth Network of Minas Gerais (TNMG). The cohort is linked to public mortality databases. Patients over 16 years old with a valid ECG performed from 2010 to 2017 were included. Clinical data were self-reported. In a 15% stratified sample of the original cohort (CODE-15) an ECG was labelled “normal” according to conventional clinical reporting and based on automated interval measurements (10).

#### (ii) ELSA-Brasil Cohort

ELSA-Brasil is a cohort study of 15,105 Brazilian public servants, aged 35 to 74 at enrolment. All active or retired employees of six participating institutions were eligible for the study. The full inclusion criteria and protocol have been previously described (11). 13,739 subjects had ECG and outcome data available for analysis.

#### (iii) The SaMi-Trop Cohort

The SaMi-Trop cohort is a prospective cohort of 1,631 patients with chronic Chagas cardiomyopathy and has been previously described in detail (12). Briefly, the inclusion criteria were: (1) self-reported Chagas disease; (2) aged 19 years or more. Digital ECGs were performed in 2011-2012 by TNMG. 83% of this cohort had abnormal ECGs (13).

#### (iv) The UK Biobank Cohort

The UK Biobank is longitudinal study of over 500,000 volunteers aged 40-69 at the time of enrolment in 2006-2010 (14). At baseline assessment participants provided information on health and lifestyle via questionnaire, had physical measures taken (including height, weight, and blood pressure) and donated samples of blood urine and saliva. A subgroup of participants were invited back for subsequent visits for additional investigations, including for detailed studies including cardiac magnetic resonance imaging (MRI), brain MRI and digital ECGs. 42,386 subjects with digital ECGs taken at the instance 2 visit were available for analysis. There is evidence of healthy volunteer selection bias (15). Outcomes were linked to cancer and death registry data, hospital admissions and primary care records. Major adverse cardiovascular events (MACE) was selected as the primary endpoint for this dataset given the low mortality rate in this healthy population and the additional data available. MACE was defined as: heart attack, stroke, heart failure diagnosis and all cause death. Detailed phenotyping using the cardiac MRI data has been previously described (16, 17) .

#### (v) The Whitehall II Cohort

The Whitehall II cohort has been previously described (18). Briefly, British civil servants were enrolled into this cohort and had repeated calls for medical investigations. Between 2007 and 2009 participants (n=5,066) had digital 12-lead ECGs performed, for 5 minutes. The first 10s of this recording was considered in our analysis. Causes of death were ascertained based on the information contained in the death certificate. The novel ECG parameters micro-QRS fragmentation and QRS-T angle have been previous applied to this cohort, the same values were used in our analysis (19).

#### (vi) The BIDMC cohort

The BIDMC cohort is a dataset comprised of routinely collected data from Beth Israel Deaconess Medical Center, Boston, USA. Subject over 16 years old with a valid ECG performed from 2000 to 2023 were included. ECGs were linked to mortality records. The first ECG recorded per subject was used in this analysis. 188,972 subjects were available for analysis.

### ECG pre-processing

12-lead ECGs were pre-processed by removing the baseline drift and re-sampling to 400hz. Zero padding resulted in a signal with 4096 samples for each lead for a 10s recording, that is used as input to the neural network model.

### Neural network-derived ECG features from CODE dataset

The CODE dataset was previously used to train a CNN to detect six common ECG abnormalities (first-degree AV block, right bundle branch block (RBBB), LBBB, sinus bradycardia, atrial fibrillation (AF) and sinus tachycardia), as previously described (8). This model is referred to as CODE-CNN. The Keras framework with a TensorFlow backend was used for neural network training and inference (20, 21).

Transfer learning can be used to repurpose a machine learning model trained for one task to another. We hypothesised the ECG features learned by the CODE-CNN could be used to cluster subjects into clinically meaningful phenogroups. In order to extract features of the CODE-CNN, the final classification layer of the model was removed (**Figure 1**) and the output of 5120 NN-derived ECG features is used to cluster the subjects.

**Figure 1.**
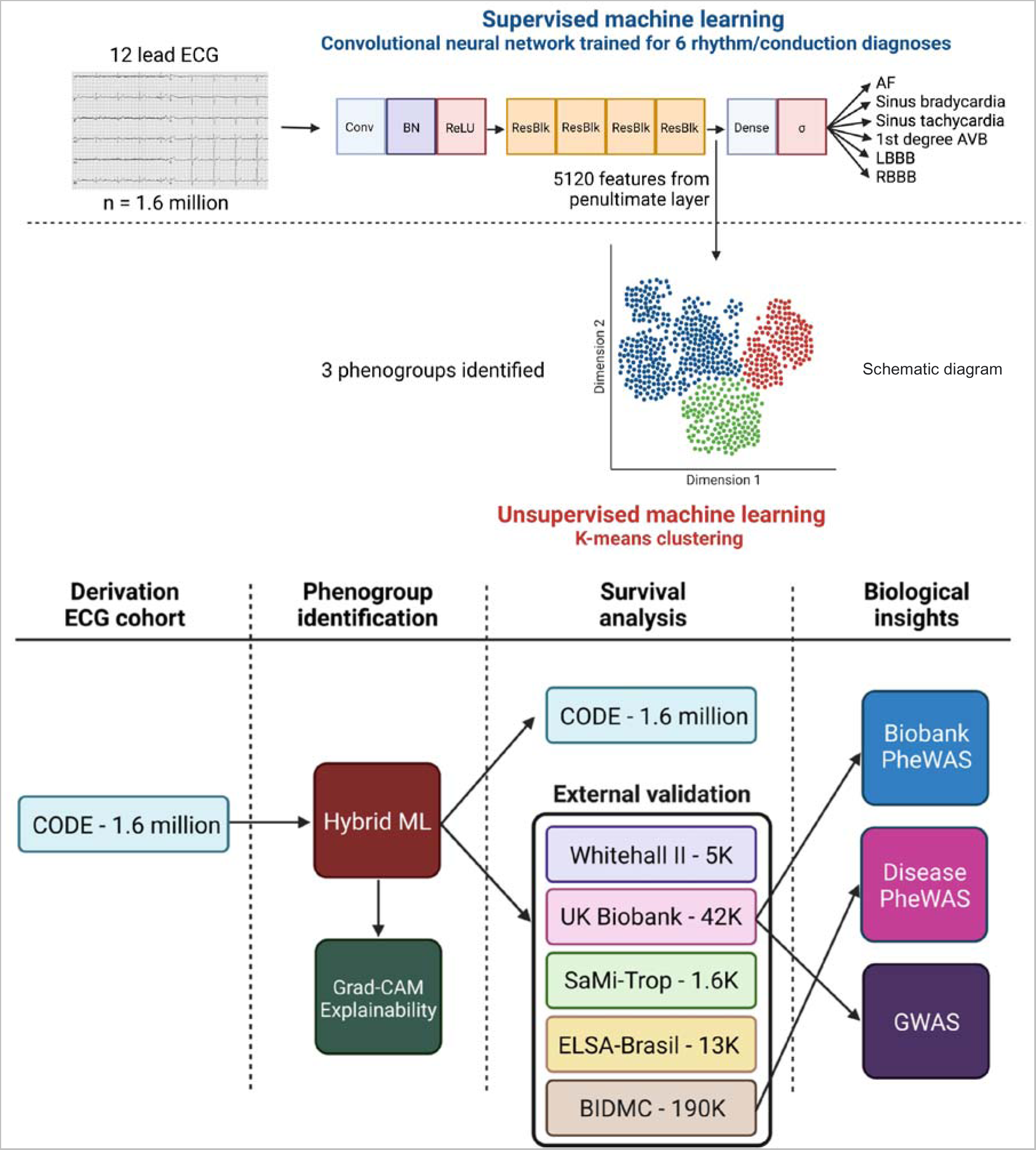
Data analysis pipeline: (A) Hybrid machine learning approach with combination of supervised and unsupervised machine learning to use NN-derived ECG features to identify phenogroups from the 12-lead ECG. 5120 NN-derived ECG features was extracted from the penultimate layer of the supervised model. K-means clustering was then applied to identify 3 phenogroups. (B) Flow of data and analyses performed in this study. AF: atrial fibrillation, AVB: atrioventricular block, LBBB: left bundle branch block, RBBB: right bundle branch block, ECG: electrocardiogram, ML: machine learning, Grad-CAM: Gradient-weighted Class Activation Mapping, PheWAS: phenome-wide association study, GWAS: genome-wide association study, MR: mendelian randomisation, CODE: Clinical Outcomes in Digital Electrocardiography, ELSA-Brasil: Brazilian Longitudinal Study of Adult Health, SaMi-TROP: São Paulo-Minas Gerais Tropical Medicine Research Center, BIDMC: Beth Israel Deaconess Medical Center.

### Unsupervised machine learning for clustering into phenogroups

In order to derive phenogroups using the 5120 features extracted using the CODE-CNN model, K-means clustering was applied in the CODE dataset. k = 3 was selected using the elbow method (22). The sklearn package in Python was used to perform K-means clustering (23). In order to determine cluster assignment in the external validation cohorts, the 5120 features were extracted using CODE-CNN as described above for each ECG. The K-means model (trained on the NN-derived ECG features from the CODE dataset) was then used to identify the nearest cluster centre and therefore determine the cluster assignment for each ECG. There was no retraining or calibration performed when applying the CODE-CNN and K-means models to the external validation datasets.

### Survival analysis in derivation and external validation cohorts

Survival analysis for the three phenogroups was first performed for the CODE dataset, and then in the four external validation datasets (UK Biobank, Whitehall II, ELSA Brasil, and SaMi-Trop). Survival analysis was repeated after excluding subjects with the six diagnoses identified by the CODE-CNN and also considering only physician-adjudicated normal ECGs in the CODE dataset. Descriptive statistics are displayed as medians (interquartile ranges) for continuous variables and numbers (percentages) for categorical variables. Kaplan-Meier plots were used to display cumulative mortality. The log rank test was used to compare survival curves. Cox proportional hazards regression modelling was used to estimate hazard ratios for mortality while correcting for other known variables. Recent work suggests virtually all real-world clinical datasets will violate the proportional hazards assumptions if sufficiently powered and that statistical tests for the proportional hazards assumption may be unnecessary (24). In line with these recommendations, the proportional hazards assumption was not evaluated and the hazard ratio from our Cox models should be interpreted as a weighted average of the true hazard ratios over the follow-up period. The adjusted variables differed in each dataset depending on the data available. Statistical analyses were performed with R 4.0.0 statistical package (R Core Team, Vienna, Austria) or Python (version 3.9).

### Phenome-wide association study

To explore more detailed phenogroup associations, we performed phenome-wide association studies (PheWAS). We used two PheWAS approaches, firstly we used a Disease PheWAS to explore the association of ECG phenogroup with incident diseases and treatments. Using the BIDMC dataset we converted International Classification of Diseases (ICD) 9 and 10 codes into Phecodes as previously described (25). In order to remove prevalent disease, we removed any incident codes if ICD codes prior to the ECG also existed. Phecodes with under 20 cases were excluded. Logistic regression was performed to investigate the association between ECG phenogroup and incident disease. The second approach, Biobank PheWAS, used the UK Biobank that contains data from over 3000 phenotypes derived from patient measurements, surveys, and investigations. Univariate correlation was performed to investigate the association between ECG phenogroup and phenotypes. Highly correlated features (>95%), features with >90% missing data or with >95% of exactly the same value were discarded. We adjusted for multiple testing using Bonferroni correction. Given the very small number of subjects in phenogroup C, subjects in phenogroup C were excluded from these analyses. Effects of age and sex were regressed out of the ECG phenogroup variable. PheWAS analyses were performed in Python (version 3.9).

### Genome-wide association study

To identify genetic associations with the ECG phenogroups, we performed a genome-wide association study (GWAS). Subjects in phenogroup C were excluded from this analysis, due to the small number of subjects, as described above for PheWAS. Using linear regression, a binary phenogroup variable was adjusted for the following covariates: age at imaging visit, sex, height, body mass index (BMI), imaging assessment centre, the first 10 genetic principal components, and genotyping array batch. Standard quality control was undertaken. Included single nucleotide polymorphisms (SNPs) had a minor allele frequency (MAF) >0.1% and an imputation INFO score of >0.4. Unrelated individuals of genetically-determined European ancestry were included as previously described (26). The GWAS was undertaken using FastGWA mixed linear model association analysis through the Genome-wide Complex Trait Analysis software using a genetic relationship matrix (GRM) to adjust for population structure (27). The Manhattan plots depict the nearest gene. Top SNPs were identified that had a P-value <5×10^-8^. Associations of the detected SNPs were evaluated using the NHGRI-EBI GWAS Catalog, PhenoScanner, GTEx, and GeneAtlas UK Biobank PheWAS browser.

### Model explainability

In order to understand the elements of the ECG contributing most significantly to phenogroup determination we modified a commonly used technique in computer vision, Gradient-weighted Class Activation Mapping (Grad-CAM) (28). A signal importance map for a given input of a 12-lead ECG signal was computed. First, the final layer of the original trained CNN is removed, and a new layer with a fixed operation is added. This layer takes the 5120 features from the trained CNN and computes its Euclidean distance to the centre of three phenogroup clusters, which results in three distance scores. Using softmin, these distance scores are mapped to probability scores. Using Grad-CAM, for a given input, we compute the gradient of weights of the first convolution layer with respect to the probability score for a specified phenogroup (e.g., phenogroup A). The output of the first layer for a given input is then multiplied by an average gradient of respective channels, and finally, the positive values are taken as the importance score for the saliency map. To extract the generalised saliency map for each phenogroup, 1000 ECGs from the centre of the three phenogroup clusters (approximately equidistant from all three cluster centroids) were taken. After computing saliency maps, each 10s ECG was averaged to one beat per lead by aligning beats with R-peaks. Similarly, the respective saliency map of each 10s was averaged to a single beat. A generalised saliency map for a cluster was computed by averaging (median) all the 1000 saliency maps. While plotting the generalised saliency map, single beats of all the 1000 ECGs were also averaged (mean) to one beat per lead.

## Results

An overview of the analysis pipeline is provided in **Figure 1**.

### Derivation cohort

Using the first recorded ECG per patient, 1,558,421 ECGs in the CODE dataset were available. Patients were followed up for a mean of 3.68±1.87 years. 52,127 (3.3%) subjects died during the follow-up period. Using a hybrid ML pipeline (**Figure 1**), three phenogroups were identified.

### Survival analysis

Our survival analysis demonstrated the 3 phenogroups had different mortality profiles (**Figure 2A**). Phenogroup B had a 2.57-fold higher risk of mortality compared to phenogroup A, while phenogroup C had a 15% lower risk (phenogroup B vs A, hazard ratio (HR) 2.57, 95% CI 2.51-2.63, p<0.001; phenogroup C vs A, HR 0.85, 95% CI 0.82-0.87, p<0.001). The prognostic significance of the phenogroups was retained, after adjusting for known covariates (age, gender, known cardiovascular comorbidities and the six ECG diagnoses on which the neural network was trained). In these adjusted analyses, phenogroup B (highest mortality) had a 1.2-fold increase in long-term mortality compared to phenogroup A (HR 1.20, 95% CI 1.17-1.23, p < 0.0001).

**Figure 2.**
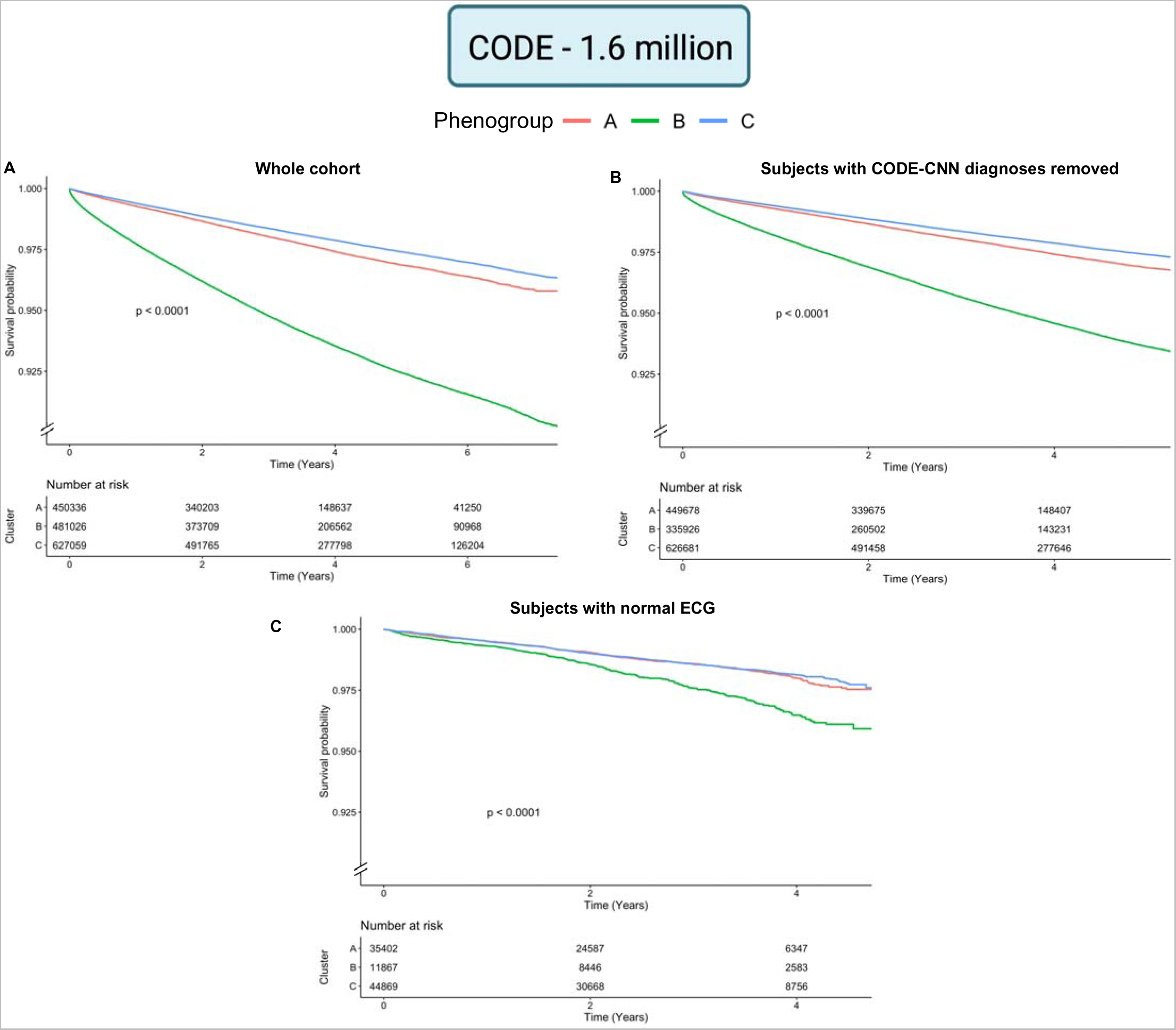
Survival analysis in the derivation dataset: The 3 phenogroups have prognostic significance, with phenogroup B having a markedly worse prognosis. Data are shown for: (A) whole Clinical Outcomes in Digital Electrocardiography (CODE) cohort, (B) CODE cohort with removal of subjects with any of the following diagnoses on the ECG: 1st degree atrioventricular block, right bundle branch block, left bundle branch block, sinus tachycardia, sinus bradycardia and atrial fibrillation. (C) Subset of the CODE cohort with normal ECGs.

We then evaluated the performance of the phenogroups in the group of patients without any of the six diagnoses for which CODE-CNN was originally trained. Importantly, the predictive ability of the phenogroups was retained in this group (n=1,412,285, **Figure 2B**). We also analysed a subgroup of patients with physician-adjudicated normal ECGs (n = 92,138). In this analysis, phenogroup B retained prognostic significance (phenogroup B vs A, HR 1.65, 95% CI 1.43-1.92, p < 0.0001) (**Figure 2C**).

### External validation

We then externally validated our findings in five diverse cohorts. There were 5,066 subjects available for analysis in the Whitehall II study. For this analysis events were censored at 5 years of follow-up. 163 (3.22%) subjects died during this period. In the UK Biobank, there were 42,386 subjects available for analysis. Patients were censored at the time of their first event and 411 (0.97%) subjects died during the follow-up period of 3.73±1.57 years. Given the comparatively significantly lower mortality rate and the additional data available in the UK Biobank, the primary endpoint for this dataset was specified as MACE (definition described methods). 967 (2.3%) of subjects had a MACE event during follow-up. The ELSA-Brasil cohort had 13,739 subjects available for analysis. Mean follow-up was 9.35±1.28 years and 599 (4.4%) subjects died during follow up. The SaMi-Trop cohort had 1,631 subjects available for analysis. Mean follow-up period was 2.08±0.39 years and 104 (6.38%) subjects died during the follow-up period. Lastly, the BIDMC cohort consists of a secondary care population. 188,972 subjects were available for analysis. Mean follow-up period was 5.46±5.81 years and 34,851 (18.4%) subjects died during follow-up.

The prognostic significance of the phenogroups was demonstrated in all of the external cohorts (**Figure 3**). In the volunteer cohorts (Whitehall II, UK Biobank and ELSA-Brasil), phenogroup C had very limited representation and therefore was excluded from analysis. This may be due to the relatively selected population enrolled into these cohorts. Cox regression models adjusted for known covariates showed the Phenogroup B retained its prognostic significance in the CODE, UK Biobank, Whitehall II and BIDMC cohorts, but not ELSA-Brasil. Importantly, this included adjustment for standard ECG parameters in the UK Biobank and Whitehall II cohorts.

**Figure 3.**
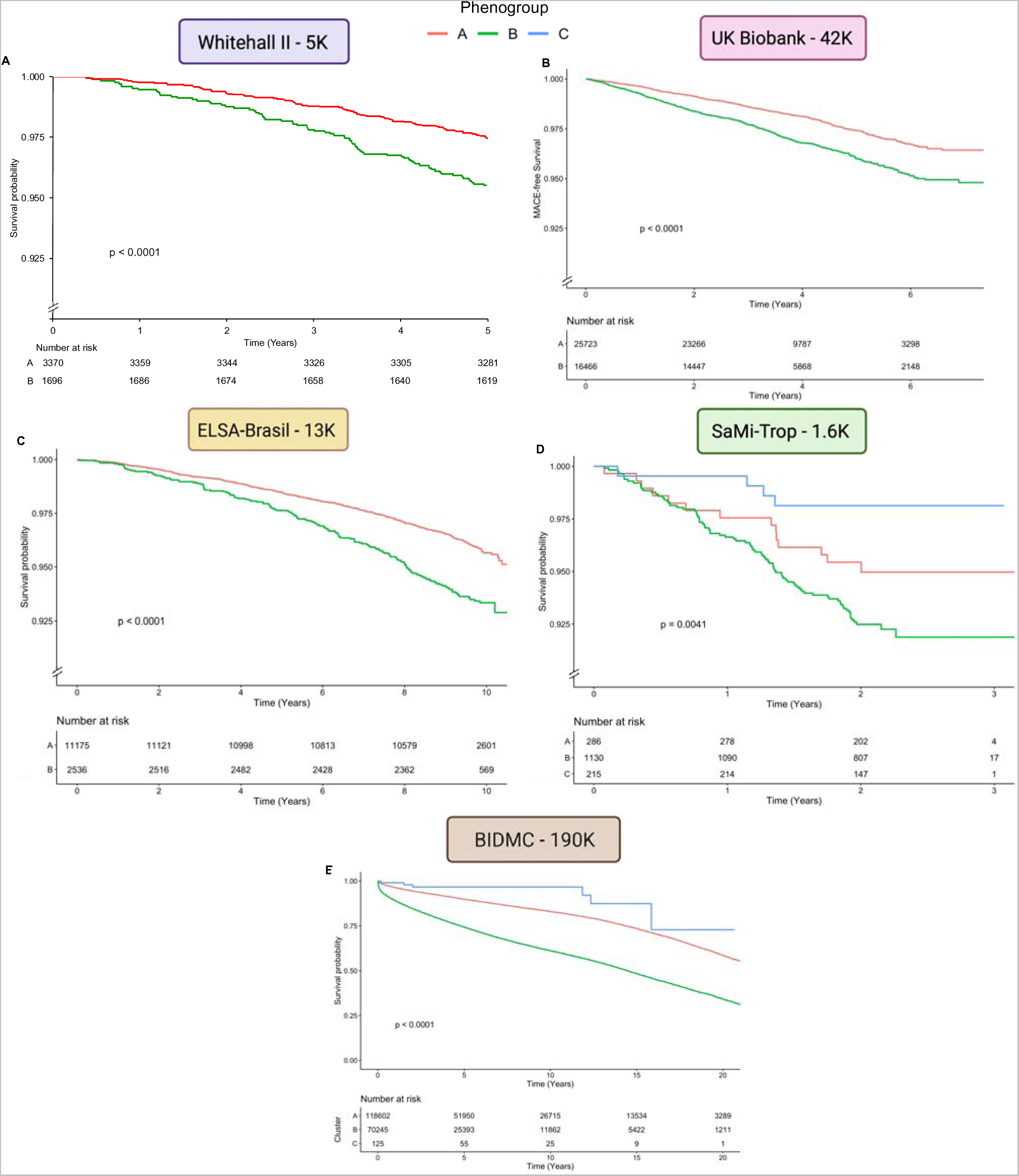
Survival analysis in the five external validation datasets. In the volunteer populations (Whitehall II, UK Biobank and Brazilian Longitudinal Study of Adult Health (ELSA-Brasil)) phenogroup C has very few subjects and therefore is excluded. Phenogroup B has a significantly higher event rate. (A) Whitehall II cohort (B) UK Biobank, survival free of major adverse cardiovascular events is depicted (C) ELSA-Brasil cohort, (D) São Paulo-Minas Gerais Tropical Medicine Research Center (SaMi-TROP) cohort, (E) Beth Israel Deaconess Medical Center (BIDMC) cohort.

To investigate the prognostic significance of the phenogroups in a more specific disease context, the SaMi-Trop cohort was analysed. In contrast to the volunteer cohorts described above (who would be expected to largely have relatively normal ECGs), most patients in the SaMi-Trop cohort had abnormal ECGs (13). In this cohort of patients with chronic Chagas cardiomyopathy, the three phenogroups had marked prognostic significance (**Figure 3D**).

We went on to perform survival analysis on the subset of ECGs without any of the original rhythm/conduction diagnoses for which CODE-CNN was trained. Phenogroup B had a statistically significant higher mortality in the UK Biobank, Whitehall II and BIDMC cohorts. Phenogroup B trended towards higher mortality but was non-significant for SaMi-Trop (p = 0.05) and ELSA-Brasil (p = 0.11).

### Phenome-wide association study

In order to evaluate phenotypic associations with the ECG phenogroups, we performed PheWAS using two approaches. Firstly, we evaluated incident disease in the BIDMC cohort (**Figure 4A**). We found the high-risk phenogroup (Phenogroup B) was associated with a significantly higher rate of future atrial fibrillation (OR 2.89, p < 0.00001), ischaemic heart disease (OR 1.44, p < 0.00001), atrioventricular block (OR 4.88, p < 0.00001), cardiomyopathy (OR 2.04, p < 0.00001), ventricular tachycardia (OR 2.00, p < 0.00001) and cardiac arrest (OR 6.68, p < 0.00001). Secondly we investigated phenotypes in the UK Biobank. The non-ECG phenotypes included broad categories of blood biomarkers, cardiac MRI features, family history, geographical factors, health outcomes, imaging parameters, lifestyle factors, physical measures, reproductive factors, and socioeconomic factors. **Figure 4B** shows the Manhattan plot of the univariate correlation P-values and correlation coefficients between ECG phenogroup and non-ECG phenotypes. 512 of 3142 comparisons reached the Bonferroni threshold for significance. The most significant PheWAS associations were in the cardiac MRI and physical measures categories. These include left ventricular (LV) and right ventricular (RV) end-systolic volume (ESV) and end diastolic volume (EDV), right atrial volume and LV stroke volume (SV) which positively correlated with phenogroup B (higher MACE event rate). Left ventricular ejection fraction (LVEF), right ventricular ejection fraction (RVEF) and measures of left ventricular strain were negatively correlated. Significant physical measures that were negatively correlated with phenogroup B: heart rate and arterial stiffness measures, systolic and diastolic blood pressure (BP), and maximum heart rate during a fitness test. Significant non-cardiac phenotypes included measures of bone mineral content (BMC), snoring and alcohol intake and carotid intima-media thickness (IMT), which were positively correlated with phenogroup B. The findings were similar when analysing only ECGs without any of the original rhythm/conduction diagnoses for which the neural network was trained (n=38,759).

**Figure 4.**
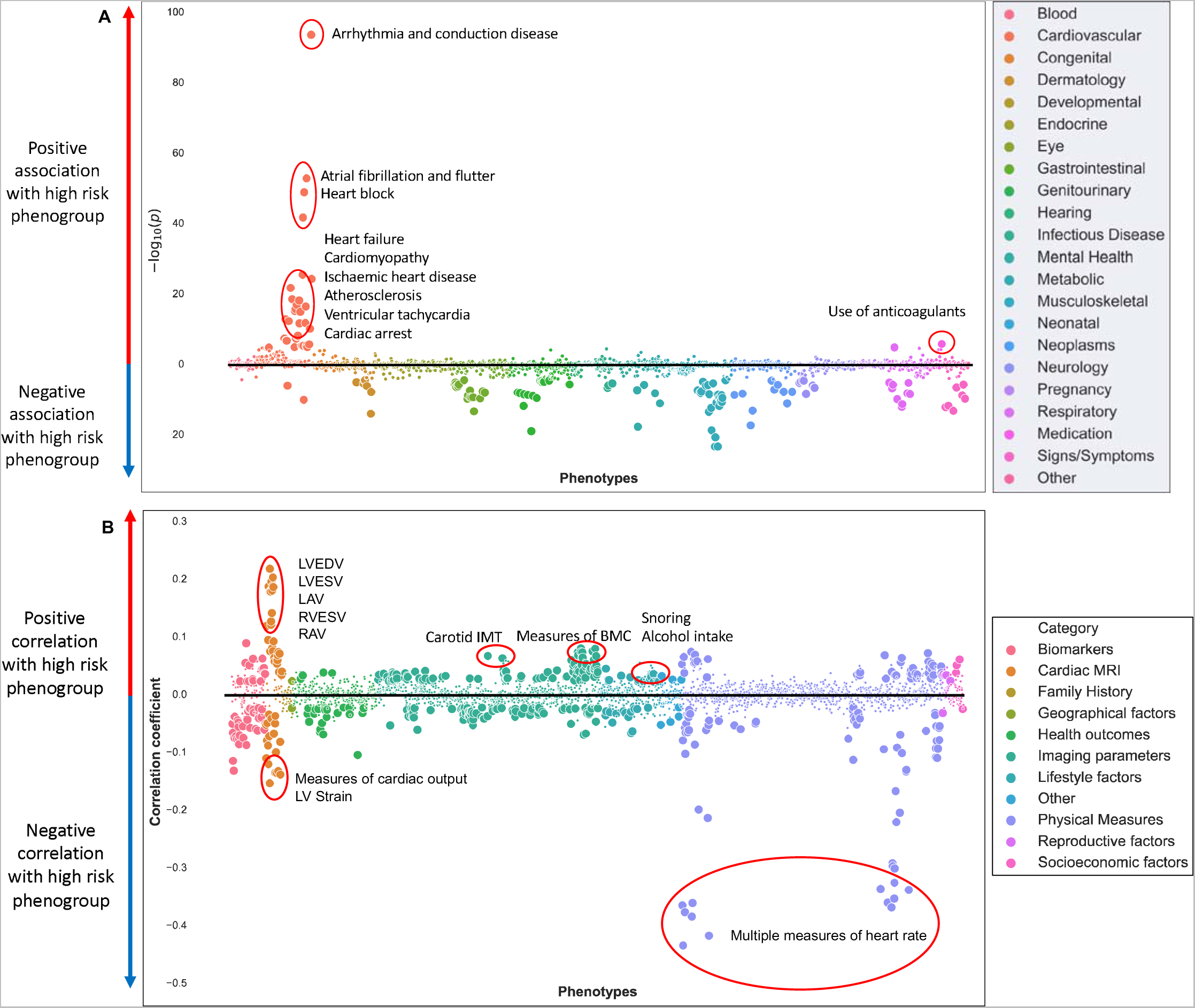
Phenome-wide association study. Manhattan plot showing negative logarithm of the univariate correlation P-value between phenotypes for BIDMC disease Phecodes, **Disease PheWAS** (A) and correlation coefficient for UK Biobank phenotypes, **Biobank PheWAS** (B). Small points depict associations not reaching statistical significance, while large points crossed the Bonferroni threshold for statistical significance. MRI: magnetic resonance imaging, EDV: end-diastolic volume, ESV: end-systolic volume, RV: right ventricle, LV: left ventricle, RAV: right atrial volume, SV: stroke volume, BMC: bone mineral content, IMT: intima-media thickness

ECG parameters also significantly correlated with phenogroup. Increased QRS duration, PR interval and QT interval, positively correlated with phenogroup B.

### Genome-wide association study

To identify genotypic associations with the phenogroups, a single-trait GWAS was conducted using 12.7 million variants in 31,119 subjects from the UK Biobank. The GWAS yielded four loci (**Figure 5**) which were confirmed through expression quantitative trait loci (eQTLs) for *Sodium Voltage-Gated Channel Alpha Subunit 10 (SCN10A), Sodium Voltage-Gated Channel Alpha Subunit 5 (SCN5A), Caveolin 1 (CAV1), and Rho GTPase Activating Protein 24 (ARHGAP24)*. The lead variant for *SCN10A* has been previously significantly associated with atrial fibrillation and flutter, and cardiac conduction (29). *CAV1* has been previously associated with QRS duration, PR interval and QT interval *while SCN5A and SCN10A* have previously well-described roles in cardiac rhythm and conduction phenotypes (29, 30, 31, 32). *ARHGAP24* has been previously associated with ECG parameters including PR interval, QRS duration and QT interval (33, 34, 35), however, our analysis has identified for the first time *ARHGAP24* as a gene associated with a prognostically significant phenogroup. The findings were similar when analysing only ECGs without any of the original rhythm/conduction diagnoses for which the neural network was trained.

**Figure 5.**
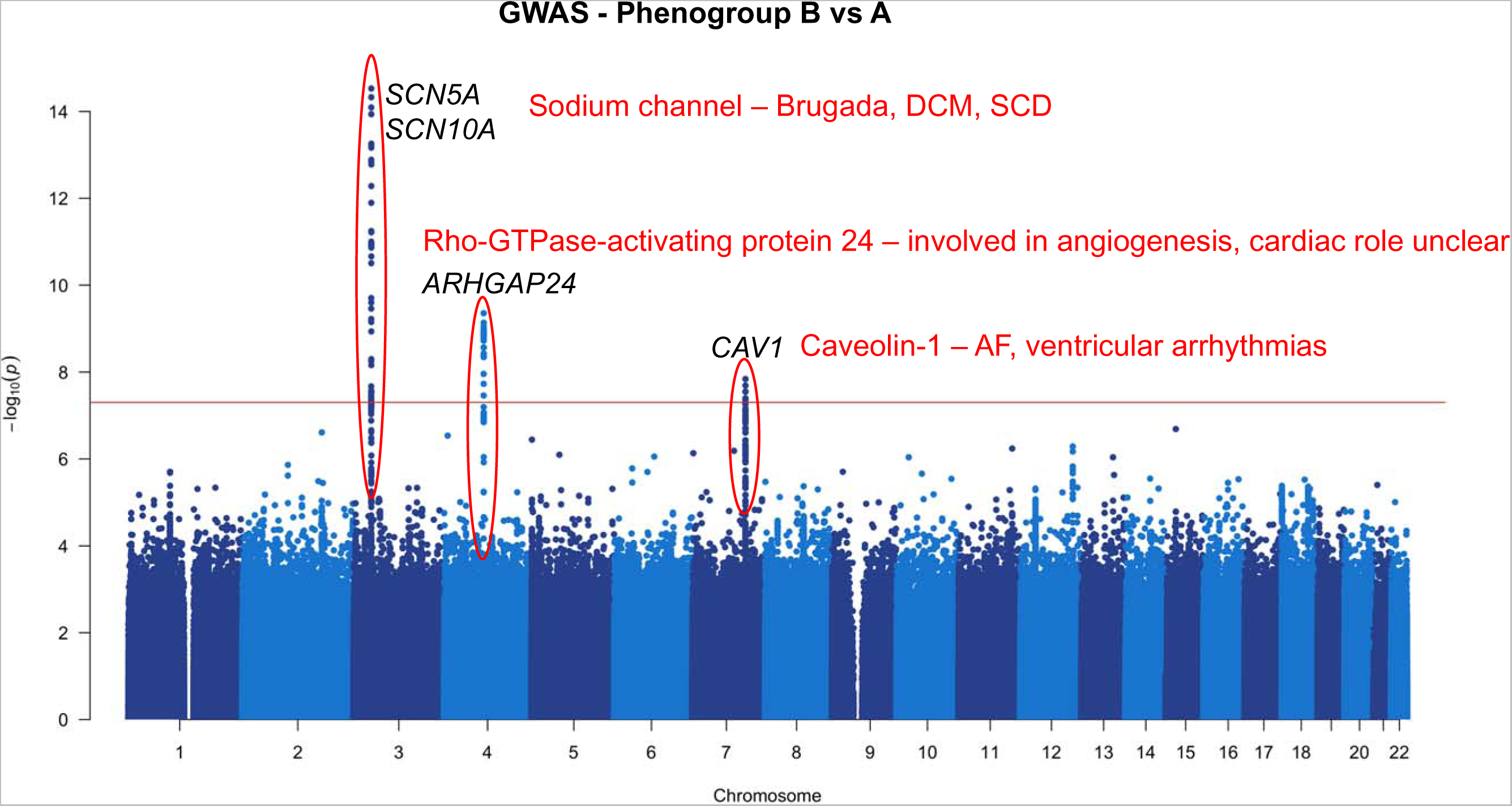
Genome-wide association study. Manhattan plots of genomic loci associated with ECG phenogroup. Nearest genes are annotated on the plot. The red line depicts the genome-wide significant threshold (P<5 x 10^-8^). DCM: dilated cardiomyopathy, SCD: sudden cardiac death, AF: atrial fibrillation

### Model explainability

In order to better understand the reasons for phenogroup classification by the hybrid ML model, we used a modified Grad-CAM approach (28). **Figure 6** shows the average saliency map for 1000 ECGs from the centre of the three phenogroup cluster centroids. The terminal part of the QRS complex and T wave were most important for identification of the high risk phenogroup (Phenogroup B).

**Figure 6.**
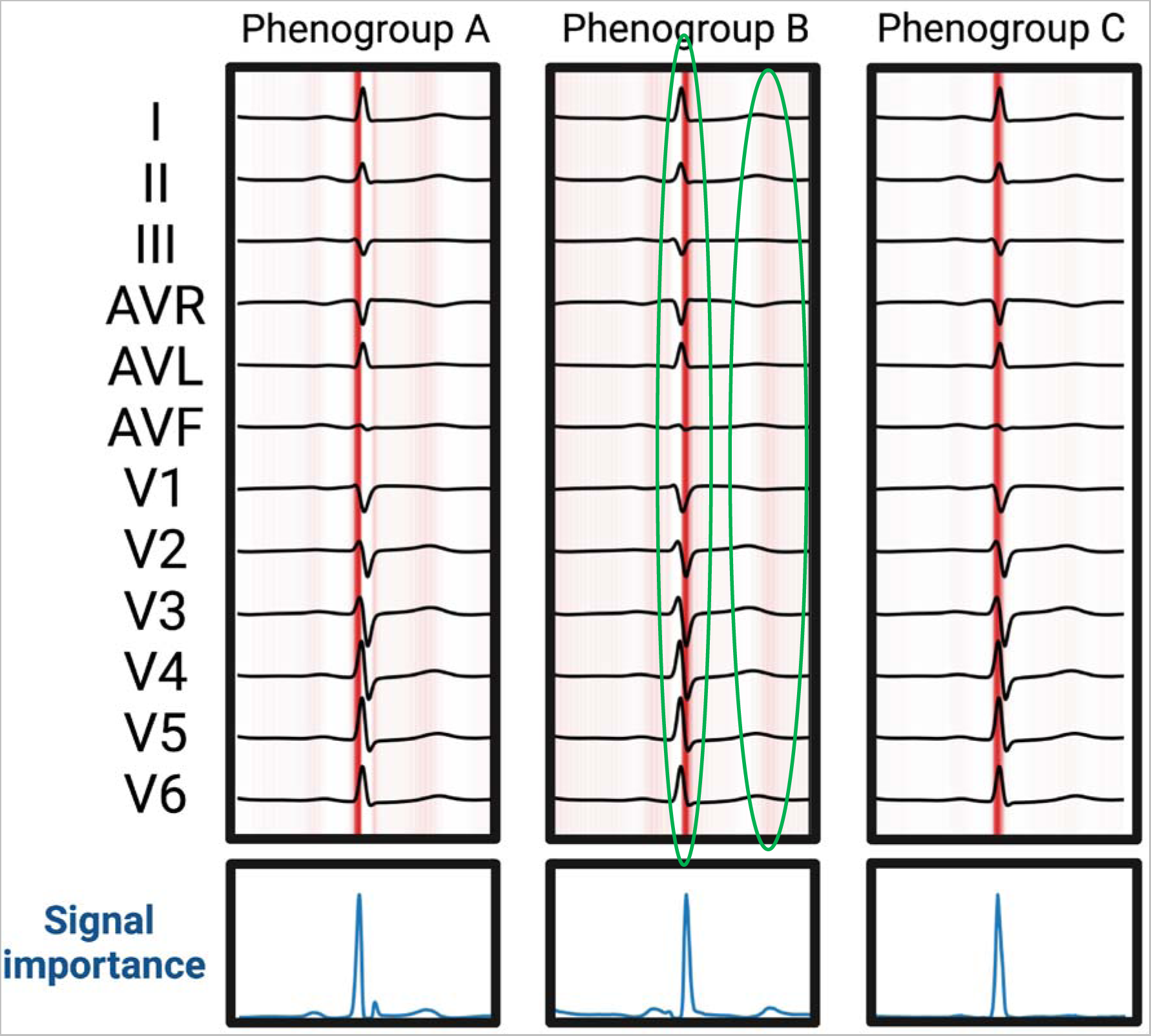
Model explainability. Gradient-weighted Class Activation Mapping (Grad-CAM) is used to generate importance maps showing the sections of the ECG signal deemed most important for phenogroup determination. Average saliency of 1000 ECGs from the centre of each cluster is shown. Areas marked with green show the terminal QRS and terminal T wave are important for identification of the high-risk phenogroup (phenogroup B).

## Discussion

In this study, we explored NN-derived ECG features to identify ECG phenogroups with prognostic significance and explored their phenotypic and genetic associations. We undertook extensive external validation across a wide range of subjects from four datasets - healthy volunteers to patients with cardiomyopathy, normal to abnormal ECGs, North and South American to European – and showed that the prognostic significance of the phenogroups was consistent across all datasets. Our analysis covers, to our knowledge, the largest ever mortality-linked analysis of a total of 1,810,215 subjects with robust and diverse external validation. Using PheWAS, we provide insight into the cardiac and non-cardiac phenotypes associated with the phenogroups. GWAS identified genetic associations primarily related to cardiac conduction and arrhythmia. MR analysis identified bidirectional pathways between the phenogroups and cardiovascular traits and outcomes. We have shown the significant potential of NN-derived ECG features, as a highly transferable and potentially universal risk marker that may be applied to a wide range of clinical contexts.

### Prognostic significance

We demonstrated that NN-derived ECG features can be used to derive phenogroups with clear prognostic significance. Although our model may in part be using existing ECG markers such as QRS duration, QRS micro-fragmentation and bundle branch block morphology (1, 2, 19), in adjusted analyses, we have shown the ECG phenogroups provide additive prognostic information beyond these.

When applied to the three volunteer cohorts (UK Biobank, Whitehall II, and ELSA-Brasil), and secondary care cohort (BIDMC) phenogroup B retained its prognostic significance despite the fact that these cohorts were demographically markedly different to the derivation cohort. In contrast to these volunteer groups, the SaMi-Trop cohort provides an insight into the prognostic significance of the ECG phenogroups in a very different context, in established disease in the form of Chagas Cardiomyopathy, where the three phenogroups show marked separation in mortality curves. This analysis shows the potential for these NN-derived features and phenogroups to be applied to a diverse range of disease specific cohorts and clinical problems, such as a broader cardiomyopathy cohort or inherited cardiac conditions such as hypertrophic cardiomyopathy.

### Biological insights

First, a Disease PheWAS in the BIDMC dataset highlighted several important incident diseases that may mediate the increased mortality seen in Phenogroup B, including arrhythmia, conduction disease, heart failure and ischaemic heart disease. Targeting screening and treatment towards these diseases is one potential strategy for reducing risk in Phenogroup B.

Second, by leveraging the extremely deep phenotyping performed in the UK Biobank, we explored the potential biological mechanisms underlying the associations of the ECG phenogroups with survival. Our Biobank PheWAS analysis has identified reduced right and left ventricular ejection fraction and abnormalities in cardiac conduction/repolarisation as potential biological mechanisms underlying the differences in prognosis of the ECG phenogroups. We report several cardiac MRI associations with the higher risk phenogroup, including lower LVEF, RVEF and left ventricular strain that have been previously associated with higher mortality (36, 37, 38) and support the association with increased heart failure and cardiomyopathy seen in the Disease PheWAS. Similarly, increased carotid IMT reflects atherosclerotic burden which supports the association with increased atherosclerosis and ischaemic heart disease seen in the Disease PheWAS (39). In contrast, higher heart rate was negatively correlated with the higher risk phenogroup, this may be due to the U-shaped relationship where both low and high heart rates are associated with increased mortality (40).

The higher risk phenogroup was associated with increased PR interval, QRS duration and QT interval. Increased QRS duration and QT interval have been previously associated with higher morality (41, 42) however increased PR interval was not previously found to be associated with increased mortality (43). Importantly, our analyses in the UK Biobank and Whitehall II cohorts have shown that ECG phenogroup adds prognostic significance beyond basic ECG parameters.

Grad-CAM highlighted the terminal QRS complex, which may represent conduction slowing, and the terminal T wave, which may represent repolarisation heterogeneity as important factors in identifying the high risk phenogroup. These factors have been previously related to adverse prognosis, which provides further reassurance to the biological relevance of our findings (44, 45).

### Genome wide-association study

Phenogroups identified from NN-derived ECG features had significant genetic correlates that provide a plausible mechanistic pathway for the difference in prognosis between phenogroups. The lead variants identified here are in genes with established roles in cardiac electrophysiology: *SCN5A, SCN10A and CAV1* (29, 30, 31, 32). *SCN5A* and *SCN10A* encode the sodium channels Nav1.5 and Nav1.8 respectively. Nav1.5 is responsible for the initiation of the cardiac action potential via the inward sodium current. Mutations in *SCN5A* and *SCN10A* have been identified as a cause of inherited syndromes such as Brugada syndrome (46, 47) and dilated cardiomyopathy (48). A role in heart failure and sudden death in ischaemic heart disease has also been described (48, 49). *CAV1* encodes caveolin-1 which is a cytoplasmic membrane-anchored scaffolding protein that co-localises to the atria and has been linked to atrial fibrillation, ECG parameters and ventricular arrhythmias (32, 50, 51). *ARHGAP24* encodes Rho-GTPase-activating protein 24 which is a key regulator of angiogenesis and is involved in cell polarity, cell morphology and cytoskeletal organization, but does not have an established role in the heart (52). The association between *ARHGAP24* and ECG parameters has been previously described, however the mechanism is unclear (33, 34, 35). In our study, we have shown a novel association between *ARHGAP24* and a prognostically significant ECG phenogroup. Our findings suggest that propensity to conduction system disease and arrhythmia may lie on the causal pathway between these variants and the increased mortality seen in phenogroup B.

### Comparison to previous machine learning approaches for ECGs

Other studies have used supervised machine learning to predict mortality (53, 54). This approach broadly involves mortality labels being provided to the machine learning model, which can then ‘learn’ the important features predictive of mortality. In contrast, an unsupervised approach involves the machine learning model clustering subjects based on features without knowledge of the mortality label. Given the ability of unsupervised or self-supervised models to ‘learn’ from massive amounts of data without knowledge of outcome labels, there is significant potential to apply these methods to clinical data, where outcome labels may be lacking.

Unsupervised machine learning has also previously been applied to ECGs for various applications. Recent studies have used a variational auto-encoder to evaluate a median ECG signal and compress it to 32 factors, these were then used to perform classification tasks using a supervised machine learning model or traditional statistics (55, 56). In this case, unsupervised machine learning was used only for feature extraction compared to our novel approach that successfully identifies clinically relevant phenogroups with no mortality labels used in model training. In contrast, Feeny et al applied unsupervised machine learning to the QRS waveforms themselves in patients who had cardiac resynchronisation therapy (CRT) devices implanted for heart failure and identified two prognostically important subgroups, with significance beyond the commonly used parameters of QRS duration and left bundle branch block. Their method of feature extraction used principal component analysis, compared to the deep learning approach used in our study. Our deep learning approach has the potential to identify more complex, non-linear, relationships between elements of the ECG.

### Potential clinical applications of NN-derived ECG features

Using a hybrid machine learning approach, we have described for the first time a novel method to identify high-risk subjects from the ECG alone. Through robust and diverse external validation, we have demonstrated the relevance of the ECG phenogroups in a wide range of clinical contexts. The addition of ECG phenogroup may significantly improve the performance of risk prediction algorithms, and therefore aid management decisions.

NN-derived ECG features could potentially be applied to risk stratification of a general population to identify high-risk individuals for more intensive investigation and follow-up and lower risk individuals who may be reassured. In disease-specific cohorts such as heart failure and cardiomyopathy, NN-derived ECG features may be applied to assess risk and guide more intensive medical therapy, implantable devices, or transplantation consideration. These potential applications however require further evaluation in the appropriate populations, in prospective studies.

### Limitations

We have described the novel application of NN-derived ECG features for risk prediction, and externally validated our findings in a diverse population. The volunteer cohorts of the UK Biobank, Whitehall II and ELSA-Brasil are selected populations. This may explain the absence of phenogroup C in these cohorts. This however does not detract from the phenogroup performance in these cohorts and in fact strengthens the findings given the diverse groups in which they were tested.

Detailed explainability for deep learning models is challenging. Although some information can be obtained through importance maps, we are unable to provide detailed understanding into the reasons for phenogroup classification. The Biobank PheWAS and GWAS analyses were performed in a population with predominantly European ancestry (UK Biobank) and findings therefore may not be generalisable globally.

## Conclusion

We describe the use of NN-derived ECG features, derived from an established AI-ECG model trained to identify 6 common diagnoses, to identify prognostically-significant phenogroups from the 12-lead ECG. We explored the biological basis underlying the difference in prognosis between the phenogroups, and identified phenotypic and genotypic associations through PheWAS and GWAS. We validated our findings in five external datasets across two continents and diverse patient populations. NN-derived ECG features have important applications beyond the original model from which they are derived and may be transferable and applicable for risk prediction in a wide range of settings, in addition to mortality prediction.

## Data availability

SaMi-Trop cohort was made openly available (https://doi.org/10.5281/zenodo.4905618). The CODE-15% cohort was also made openly available (https://doi.org/10.5281/ zenodo.4916206). Restrictions apply to additional clinical information on the CODE-15% and SaMi-Trop cohorts; to the full CODE cohort, the ELSA-Brasil cohort, and the Whitehall II cohort. UK Biobank data are available upon application (http://www.ukbiobank.ac.uk/). The BIDMC dataset is restricted due to ethical limitations. Researchers affiliated to educational, or research institutions may make requests to access the datasets. Requests should be made to the corresponding author of this paper. They will be forwarded to the relevant steering committee.

